# Evolving SARS-CoV-2 variants and mutational cascades

**DOI:** 10.1101/2021.04.03.21254871

**Authors:** John M. Halley, Despoina Vokou, Georgios Pappas, Ioannis Sainis

**Affiliations:** Laboratory of Ecology, Department of Biological Applications and Technology, Faculty of Health Sciences, University of Ioannina, 45110 Ioannina, Greece; Department of Ecology, School of Biology, Aristotle University of Thessaloniki, 54124 Thessaloniki, Greece; Institute of Continuing Medical Education of Ioannina, Ioannina, Greece; Medical School, Faculty of Health Sciences, University of Ioannina, 45110, Ioannina, Greece

**Keywords:** mutation, SARS-CoV-2, hyper-exponential growth, variant of concern, SEIR model, transmission coefficient

## Abstract

The emergence of novel SARS-CoV-2 variants of concern (VOC), in late 2020, with selective transmission advantage and partial immunity escape potential, threatens a pandemic resurgence. The timing of mutational evolution and its limits are thus of paramount importance in preparedness planning. Here, we present a model predicting the pattern of epidemic growth including the emergence of variants through mutation. It is based on the SEIR (Susceptible, Exposed, Infected, Removed) model, but its equations are modifiable according to the transmission parameters of novel variants. Since more transmissible strains will drive a further increase in the number of cases, they will also lead to further novel mutations. As one cannot predict whether there is a viral mutational evolutionary limit, we model a cascade that could lead to hyper-exponential growth involving the emergence of even more transmissible mutants that could overwhelm systemic response. Our results are consistent with the timing, since the beginning of the pandemic, of the concurrent and independent emergence of the VOCs. We examine conditions that favor the expected appearance of similar variants, thus enabling better preparedness and relevant research.

## Introduction

New variants of SARS-CoV-2 have been appearing steadily throughout 2020-2021. The prospects of a mutating virus have prompted considerable expressions of concern about increases in virulence, ability to infect previously low-risk groups like children, and lower susceptibility to the neutralizing effect of both serum from convalescent individuals and antibodies emerging post vaccination. This is because the appearance of highly infectious new strains will raise the overall growth of the global pandemic unless action is taken. Of special concern has been the appearance, in late 2020, of new variants of COVID-19, with a higher reproductive number (*R*_0_) and partial immune resistance, mainly due to accumulated mutations in their spike protein. These include the UK strain (B.1.1.7/501Y.V1, August 2020), the South Africa strain (B.1.351/501Y.V2, October 2020) and the P.1 lineage identified in Brazil (P.1/501Y.V3, January 2021)(1, 2). Further variants of concern have been reported from Uganda (3), the Philippines (4) and in Northeast US (5).

Although most RNA viruses accumulate mutations as a result of their RdRp infidelity, the SARS-CoV-2 was initially thought to be less prone to mutations, since its RdRp incorporates proofreading activity (6). Unfortunately, soon after the expansion of the pandemic (February 2020), a mutation at position 314 of the ORF1b resulted in an amino acid change of the RdRp that led to an increase of subsequent mutations, as Pachetti et al (7) argued, including those on the spike (8). The ascendance of highly infective strains is an expected development in a pandemic situation since they have a selective advantage, irrespective of their pathogenicity.

An increasing rate of infections itself increases the total number of mutations providing the opportunity for the emergence of new strains of even higher infectiousness. The resulting cascade effect could cause the dynamics to develop a hyper-exponential character. Hyper-exponential growth (HEG) is essentially exponential growth with an increasing exponent. HEG is seen in the growth of the human population (9) and economy (10). It is rarely observed in nature and appears only in evolutionary systems under selection for growth. It has also appeared in some models of genetic instability in neoplasms (11). In the case of a growing pandemic, as more people become infected, the number of individual viruses does too, with it the total mutation rate, and with them the probability of emergence of a viable new variant. If this variant, in addition, has a higher reproductive number, *R*_0_, it will increase more rapidly than the original, eventually overtaking it. As a result, the next mutation of significance (conferring higher *R*_0_) is expected to derive from this new strain. We will thus observe the emergence and ascendance of higher-*R*_0_ strains. Less infectious variants, with low or negative growth rate, quickly fall behind. Now the epidemic spreads faster, with more infections, more mutations and the emergence of ever more successful variants. This cascade rapidly moves the process into a hyper-exponential pattern of growth that continues until the epidemic runs out of susceptibles and the number of infected people starts to decline.

The actual emergence and spread of the new aggressive strains shows that such mutations in SARS-CoV-2 are indeed enjoying a selective advantage. Similarly, it can suggest the onset of a process deviating from exponential growth. By using a model of epidemic growth including mutations that enable increased *R*_0_, the purpose of this study is to examine how likely it is for this process to continue further into HEG in the pandemic of COVID-19. The possible disruptive aspects of HEG, should it happen, deserve scrutiny because they would involve at least greater numbers of cases, more rapid spread, stricter lockdowns, and larger vaccination coverage.

Our approach uses a system of SEIR equations (12, 13) modified to include new variants with variable transmission rates. We model the expansion of the disease in an immunologically naïve population. Selection is by growth rate alone, so that significant mutations (leading to VOCs) are based only on the conferring of higher reproductive numbers.

## Results

In Fig. 2b, we can see the epidemic growing initially at a rate of 0.031 per day, the average growth of the COVID-19 pandemic in 2020 (14). The first mutant strains (orange curves) with greater “fitness” arise at day 180 and increase more rapidly than the initial strain. Three such order-1 variants appear before the order-2 variants (red curves) start to emerge, at day 270 (red curves), and increase still more rapidly. This is the driving force beneath the upturn of the rate of infections after day 400. Together with this, there is a massive increase in the number of new successful variants (mostly first and second order) simply because of the huge number of cases overall. Some strains with very high growth rate can be seen in Fig. 2b but as they do not reach high numbers, they do not significantly alter the overall trajectory.

**Figure 1.**
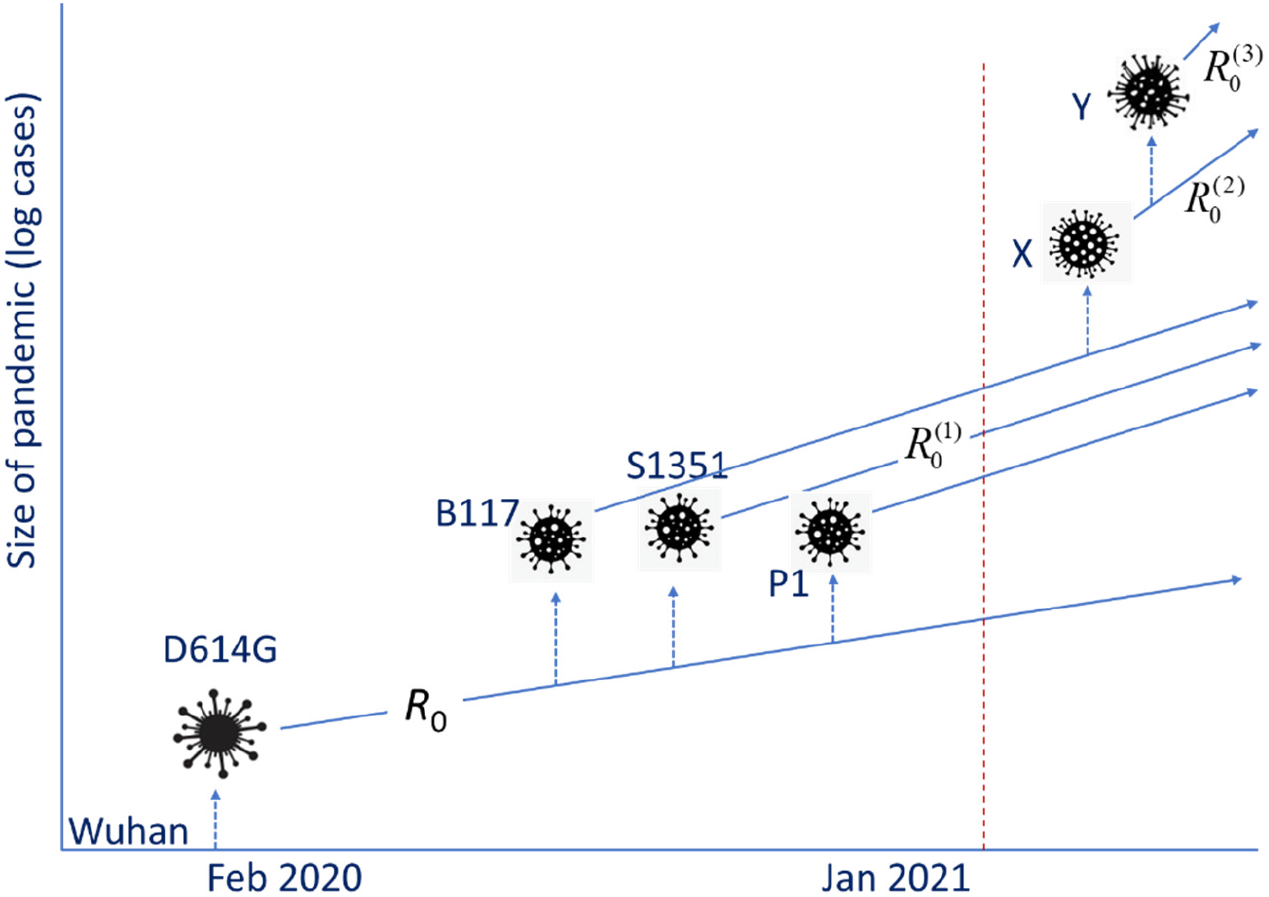
Schematic depiction of the present and a possible future scenario of the evolution of the pandemic. Dotted arrows indicate mutations, while the straight lines depict the slope of the exponential growth in log-space. The original ‘Wuhan’ variant was replaced by D614G, which has spawned in turn several successful variants with a higher reproductive number (*R*_0_^(1)^). This is the current evolution of the pandemic. Future developments (to the right of the red broken line) could include the emergence of more variants (‘X’, ‘Y’ etc.), with even greater reproductive numbers (*R*_0_^(2)^, *R*_0_^(3)^etc.)

**Figure 2.**
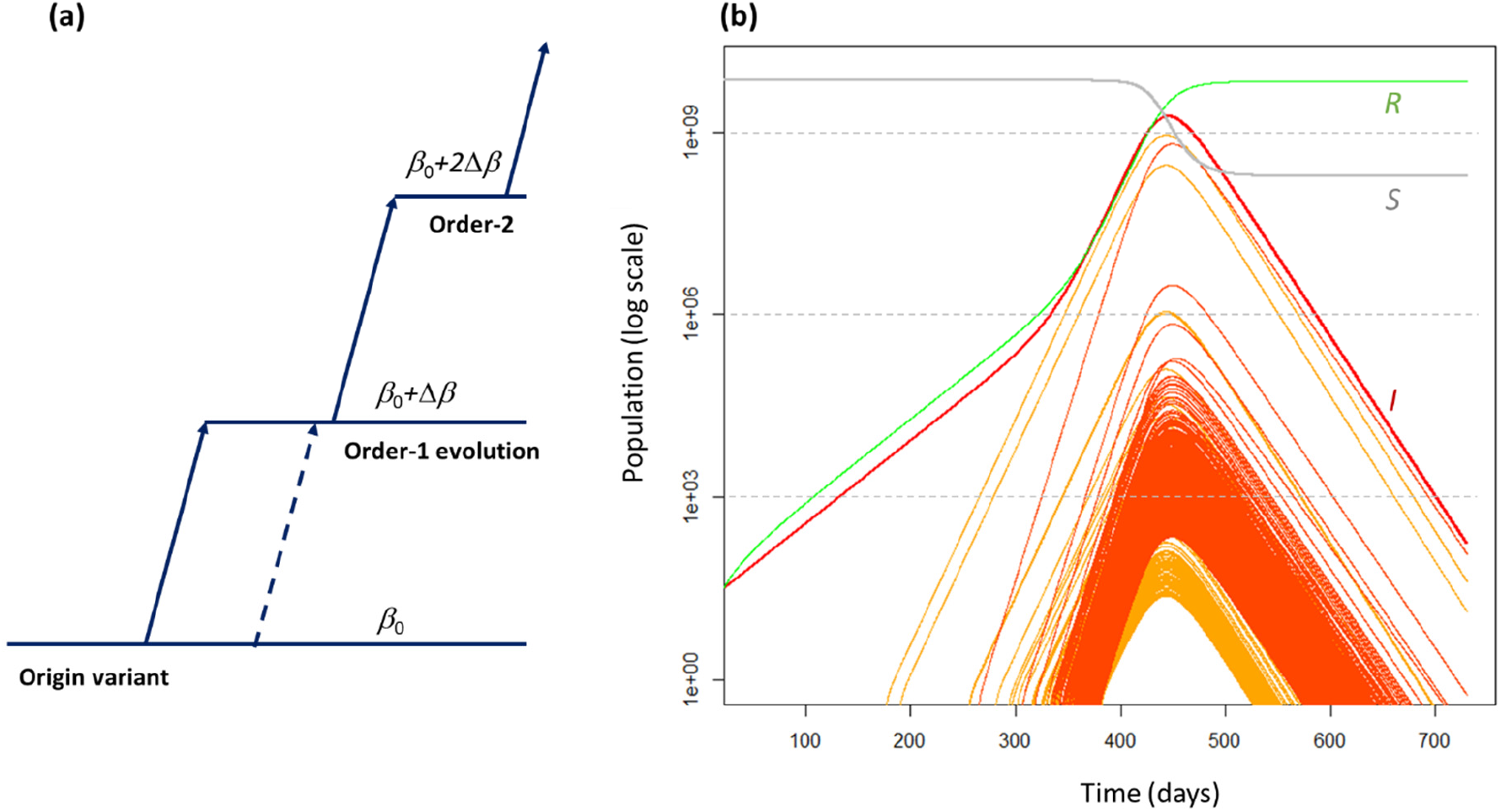
**(a)** Model of mutating SEIR system used in this paper. Each variant may increase its transmission coefficient by changing its traits, one mutation at a time. If there are *k* successful mutations, this “order-*k*” change delivers an increase *k*Δ*β* in transmission coefficient. Other mutations of the original strain can create other order-1 variants at any time (broken lines). A single mutation cannot raise the transmission rate by more than Δ*β*. Thus, a variant cannot jump directly to order-2 or higher from the original. **(b)** Solutions of modified SEIR equations (Eq. **2**): the thick red curve is the total number of infectious individuals (*I*), the green curve is the number of susceptibles (*S*), and the grey curve is the number of removed (*R*). Pre-infectious numbers are not plotted. The orange curves are the numbers of infectious people with order-1 strains of the disease while red curves are for those of order-2 or higher. The initial values of the SEIR parameters were: *E*_0_=27, *I*_0_=0, *S*=7×10^9^, *R*=0, transmission rate *β*_0_=4.57×10^−12^ per day, and Δ*β*=0.9*β*_0_. Latency time was 5.2 days and recovery time 14 days. The simulations covered a period of two years. The probability of a successful mutation is *p*=10^−5^ per infected individual. The equations were solved using the Euler method with a step size Δ*t*=0.2 days.

The probability *p* of a successful mutation determines the output of this model. There are three types of outcome. If the probability of a successful mutation is very small (*p*<10^−6^ per infected person), the overall dynamics of the pandemic are not noticeably affected, despite new strains appearing. If it is large (*p*>10^−3^), there is a strong cascade effect, with a marked curvature of the number of infectious individuals, implying sequential accelerations and HEG. Finally, for intermediate values of *p*, we tend to see the pattern of Fig. 2b, i.e. a large number of highly infectious strains dominated by a few variants.

## Discussion

The probability of a successful mutation, *p*, is crucial because it determines which of the possible outcomes (no change in the system, cascade effect or a few dominant variants) will happen. For a mutational cascade to develop, a probability greater than 10^−3^ per infected person is required. Such large values seem unlikely in the present pandemic. A more likely outcome is that a single mutation increases the *R*_0_ but without developing into a cascade of increasing *R*_0_. This happens because there is enough time for the first variant to infect most of the susceptible population but not enough time for the faster strains to reach high numbers and spawn further successful new strains. The new strain D614G that went on to replace the original Wuhan variant globally was first identified in February 2020 (15), when the total cumulative number of infected individuals was below 100,000; this would be consistent with a value of *p>*10^−5^, the value we used.

The COVID-19 pandemic represents an inadvertent experiment involving viral infection and mutation. Similar dynamics would not have been seen in other pandemics either because their expansion happened prior to the age of scientific observation or because they have been limited in their expansion (SARS-CoV-1). Interestingly, all the previously recognized RNA viral epidemic diseases with higher *R*_0_ concern viruses with smaller genomes than SARS-CoV-2. As SARS-CoV-2 has been established in the human population, in contrast to SARS-CoV-1 and MERS, the question then arises whether the accumulation of mutations associated with its larger genome might confer a greater potential for infectiousness compared to previous RNA viral diseases. Selection on the basis of growth rate alone can happen in the expansion phase of any invasive organism; this type of selection may be very different to that in an established population. For example, in the expansion of the cane toad (*Rhinella marina*), individuals tend to have longer legs on the invasion front (16), but once the population is established, shorter legs return (17). Likewise, once the “frontier” closes for SARS-CoV-2, its evolution may join the more familiar pattern in other epidemics, optimizing for long-term coexistence with its hosts.

Hyper-exponential growth, which can be understood as an accelerating exponential curve, is of interest for a number of reasons. In addition to the current concern, HEG does not arise in most biological systems but in systems involving some sort of adaptation and innovation (18). In contrast to exponential-type processes, such explosions reach a population singularity in *finite time* (10, 18). Such a model follows, as a differential equation, from the SEIR or related SIR (Susceptible, Infected, Removed) model, if mutation is included.

Our model is by necessity a simple one. We have not included spatial effects in it, nor the effects of age structure in the human population. These are factors that need to be incorporated when considering the worrying possibilities of uneven lockdown or non-homogenous vaccine coverage, that might allow intense localized viral circulation. Our model of evolution is simplified and linear: while we consider different values for mutation probability, in any simulation, the probability of successful mutation (per infected person) is constant. We have assumed that mutations of viral traits are all independent, although, in reality, they may include trade-offs. In presenting our model, we have focused on the basic patterns we are likely to see when successful mutations arise in an expanding pandemic.

Following the emergence of D614G from the original Wuhan variant in February, the arrival of the three VOCs that emerged in late 2020 may be seen as the evolution or an order-2 variant from an order-1 variant. Assuming there is no maximal viral advantage embodied in that configuration (found in different novel strains with both E484K and N501Y arising independently in different world regions), our model predicts the timing of emergence of even more problematic higher-order variants. In all runs of this model, large numbers of new strains emerged. The probability that the events will unfold similarly to Fig 2b is hard to predict because these depend on factors not included in the model, such as spatial configuration and human responses. Our findings underline the need to minimize inhomogeneous vaccine coverage since failure to do so along with other factors like “social distancing fatigue” (13) could contribute to pandemic resurgence and the possibility of HEG developing.

## Materials and Methods

Several authors have modeled the current pandemic in various places by a system of SEIR equations (12, 13). We will use this model without age or spatial structure. We assume individuals who get infected spend on average *D*’=5.2 days in an asymptomatic pre-infectious phase and then *D*=14 days upon becoming infectious before death or recovery (12, 13), although the actual infectious period may be far shorter. In 2020, the global pandemic increased from a weekly average of 26 cases day^-1^ (20^th^ January) to one of 579,000 day^-1^ (5^th^ December), equivalent to an exponential growth rate of 0.031 day^-1^, which we take as the basic growth rate. With each new infection, there is a fixed probability of a mutation causing a significantly higher growth rate (Fig. 2a). Such successful mutations happen according to a Poisson process, with fixed rate per infected person. We assume that this enters through the transmission parameter *β* in the equations, so that only the growth rate is affected. A virus with a transmission parameter *β*_0_ evolves into one with a parameter *β*_0_+Δ*β* and an associated reproductive number *R*_0_^(1)^ (order-1). Further successful mutations (order-2, order-3, etc.), with transmission rates *β*_0_+2Δ *β, β*_0_+3Δ*β*, and so on, lead to still higher reproductive numbers *R*_0_^(2)^, *R*_0_^(31)^ …, respectively. This is typical for RNA viruses that are causative agents of major diseases throughout human history and which are transmitted via respiratory droplets or aerosols, including the measles virus with a *R*_0_ about 12-18, mumps (*R*_0_ ≈10-12) and rubella (*R*_0_ ≈6-7) (19). We assume a limit for *R*_0_ to be 20, close to the highest known *R*_0_ (measles) for such viruses. We can associate these additive increments with successively adapting traits (spike protein, infectious period (20), heat resistance, etc). Our model follows each new strain that emerges from a significant mutation. We ignore strains that mutate to the same or lower *R*_0_, we do not include in the model complex mutations like blooms or super-spreader variants, and we do not make specific provisions for immuno-compromised populations that may harbor viral persistence, potentially initiating a novel variant. All calculations were performed in R (21).

### Simulation model: The SEIR with evolving transmission rate

We begin with the standard SEIR model for a single strain (19) that contains equations for the number of susceptible people (*S*), the infected hosts (*I*_0_) and the number recovered or dead (*R*), plus an equation for the number of pre-infectious people (*E*_0_), so we have:

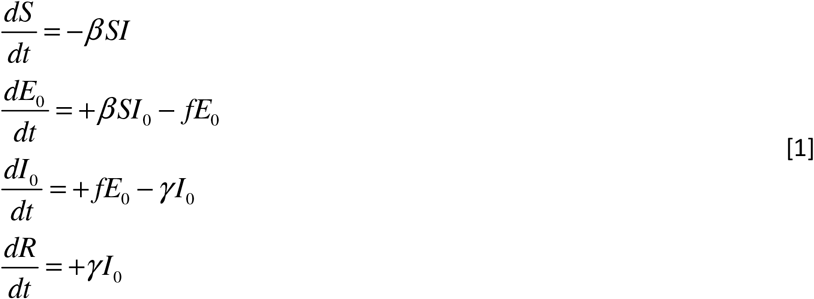

Here, *f=1/D’*, where *D’* is the latency period, *β* is the per capita transmission rate and *γ* is the recovery rate. In Eq. 1, the infected stages have a subscript ‘0’ because they refer to the original strain. There may be several strains but the susceptible and recovered/removed sub-populations are common to all strains.

In our model, each subsequent strain, *k*, has its own subpopulation of pre-infectious and infectious individuals (*E*_*k*_ and *I*_*k*_, respectively). Thus, if the total number of strains is *K*+1, we have *K*+1 pre-infectious {*E*_0_, *E*_1_, *E*_2_, …, *E*_K_} and *K*+1 infectious {*I*_0_, *I*_1_, *I*_2_, …, *I*_K_} subpopulations with reproductive rates at least as high. Thus, we now have 2*K*+4 equations in the system:

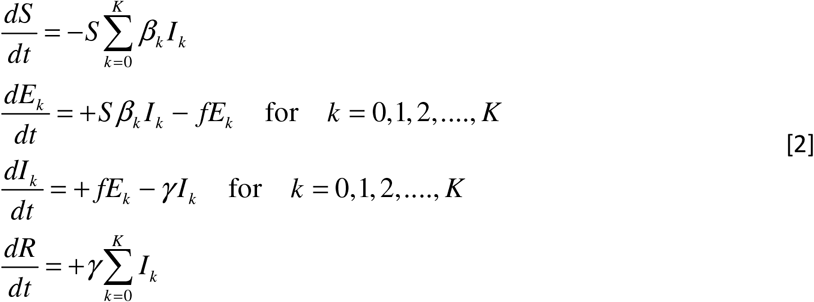

We assume that the latency time and the recovery rate are the same for all *K*+1 variants, but the transmission coefficient is assumed to change for each strain. This system of equations is solved by the Euler method using a time step of Δ*t*.

For a system with strains 0,1,2,…, *K* in circulation, the creation of a new strain “*K*+1” proceeds as follows: For each host-pathogen interaction, we assume a probability *p* of a successful mutation establishing a new strain. There are *β*_k_*I*_k_(*t*)*S*(*t*)Δ*t* such interactions in the time step of length Δ*t*, based on Eq. 1(a). Thus, the probability of a successful new mutation emerging is *p*_m_=*pβ*_k_*I*_k_(*t*)*S*(*t*)Δ*t*. In the simulations, the creation of a new strain is decided by a uniform random variable *U* on the unit interval. If *U*<*p*_*m*_, the new strain is created and the population of *E*_*K*+1_ jumps from zero to unity in the time interval [*t, t*+Δ*t*] so that *E*_*K*+1_(*t*+Δ*t*)=1. Then, the new strain has a transmission coefficient of

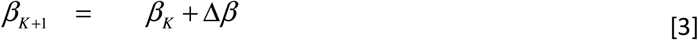

Suppose Δ*β* =0.9*β*_0_ and four variants existing, variant-0 with transmission coefficient *β*_0_ and three others, each with transmission coefficient 1.9 *β*_0_. If a mutation happens in variant-0, the result is the creation of a new strain-4, again with 1.9 *β*_0_. If, however, the mutation happens in one of the other strains, we have a new strain with transmission coefficient *β*_4_= 2.8 *β*_0_.

We assume that a significant mutation, one that increases transmission rate by more than Δ*β*, has a probability *p*, which is typically very small for an individual. For such a mutation to happen somewhere in the population, when *R* individuals have had the disease after a time Δ*t*_1_, then *pR* should have reached at least unity. *R* is found by integrating Eq. 1(d) for a variant that grows exponentially from a single infection:

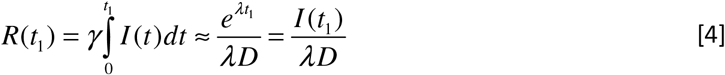

Using the requirement that *R*(*t*_1_)≈1/*p*, we can invert this to get the time-interval for the appearance of the new variant,

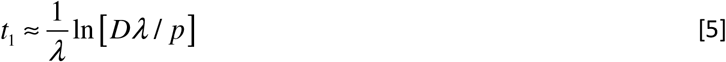

Thus, if *D*=14 days, and we assume a doubling time of 2 weeks (*λ*=0.15), using these numbers in Eq. 5, we get the first successful mutation appearing after *t*_1_=66 days, if *p*=10^−4^, or after *t*_1_=112 days, if *p*=10^−7^. Subsequent variants of the same order will occur more quickly because of the higher abundance, given the exponential growth (see Fig. 2b). However, the time for these new strains to reach sufficient abundance and start generating their own mutations may be substantial. For achieving parity with an exponentially growing variant of initial abundance *I*_0_, the new mutant with Δ*λ* greater growth rate still needs time:

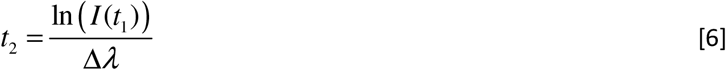

For example, the D614G variant, appearing around 20^th^ February 2020, took three months to attain dominance over the D614C form (15). If we use a figure of 52,000 (the sum of new cases over the previous 14 days) as an estimate of the number of infected people *I*(*t*_1_), then a difference of Δ*λ*=0.12 per day would bring about parity within 90 days.

### The Differential Equations of Hyper-Exponential Growth

The SIR equations, based on a simpler model that assumes that the latency period is small, may be written as follows (22):

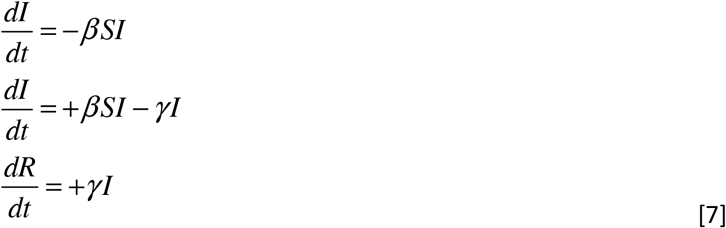

In the earliest stages of the epidemic, the number of infections is small relative to the total population, so *S≈N*. We can then solve Eq. 7(b) by solving the equation d*I*/d*t ≈*(*βN-*λ*). I≈ *λ*I*. Since *λ* is constant, the solution of this is exponential growth with rate *λ*. Note that, in the SIR model, *λ* is relSated to the reproductive number by *λ=*(*R*_0_-1)*γ*.

We assume that the per capita transmission coefficient, *β*, can be altered by successful mutations. If these are plentiful, we assume that there is a large number of successful or neutral mutations independent of each other and each leading to the same increase in *β*, and correspondingly in growth rate, *λ*. We refer to the incremental increase in *λ* as Δ*λ*. If the number of people who have had the disease is *R*, using Eq. 7(c) and the fact that the probability of a successful mutation is *p*, then the change in the number of successful mutations is proportional to *R*, i.e. Δ*λ ∝p*Δ*R*, so we can submit this into the third equation to get:

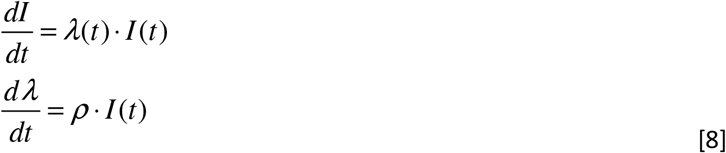

where the constant *ρ∝ p/D*.

## Data Availability

All data or code upon which this article is based will be made available on reasonable request.

## Acknowledgements

We thank Pej Rohani, Luis Borda de Agua, Paulo Borges and Jason Matthiopoulos for helpful comments.

